# Efficacy of Vitamin C in Acute Musculoskeletal Pain Management: A Double-Blind Randomized Controlled Pilot Study

**DOI:** 10.64898/2026.02.25.26347033

**Authors:** Raoul Daoust, David Williamson, Caroline Arbour, Jeffrey J Perry, Simon Berthelot, Vérilibe Huard, Patrick Archambault, Marcel Émond, Dominique Rouleau, Judy Morris, Justine Lessard, Maryse Kochoedo, Alexis Cournoyer

## Abstract

**Introduction:** Recent evidence has shown that vitamin C has analgesic properties in immediate postoperative context. However, while a clinical trial is currently underway to evaluate vitamin C for reducing opioid consumption in acute musculoskeletal (MSK) injuries emergency department (ED) patients, its direct analgesic effect in this population has not yet been established. This pilot study evaluated the feasibility of conducting a randomized placebo-controlled trial to determine the analgesic effect of vitamin C alone compared with placebo in acute MSK injured ED patients.

**Methods:** We conducted a double-blind, randomized controlled pilot trial stratified by fracture status in a tertiary care center. Adults (≥18 years) presenting to the ED with MSK injuries of ≤ 48 hours’ duration and pain intensity >3/10 were randomized to receive vitamin C 900 mg twice daily for three days or placebo. Participants completed a six-day diary (electronic or paper) and were contacted on day six to document analgesic use, treatment adherence, and pain intensity.

**Results:** Overall, 147 patients were screened; 63 (42.9%) were excluded, 24 (16.4%) refused, leaving 60 (41.1%) participants, with a consent rate of 13.0/month. Mean age (SD) was 41.8 years (14.23) and 50% were female. Lost to follow-up rate differed between participants with electronic diary (n=7; 16.7%) and participants with paper diary (n=10; 55.6%). Patients’ compliance with treatment was 97.6%. The least-squares mean difference between group A and group B in the time-weighted sum of pain intensity differences over 72 hours (SPID72) was 348.7 (95% confidence interval [CI]:-698.9 to 1396.4) for the intention-to-treat analysis and 357.6 (95%CI:-709.67 to 1424.82) for the per-protocol analysis.

**Conclusion:** This pilot study supports the feasibility of a larger randomized controlled trial on the analgesic properties of vitamin C for acute MSK injured ED patients. Strategies to reduce the missed patients and lost to follow-up rates are proposed.

**Trial registration number:** NCT06306183, ClinicalTrials.gov

## Introduction

Preclinical studies using animal models have recently shown that vitamin C (ascorbic acid) exhibits analgesic properties, providing both pain relief and opioid sparing effects in addition to its well-known antioxidant role [1, 2]. Several mechanisms have been proposed to explain these analgesic effects. One involves the attenuation of free radical production, thereby limiting oxidative injury to tissues, including neural structures [3]. Another relates to vitamin C’s essential function in collagen synthesis within bone and connective tissue, which supports tissue repair and wound healing [3, 4].

Clinical research has begun to investigate whether these biological effects translate into meaningful reductions in postoperative pain. Four meta-analyses, including two in 2025, have evaluated the impact of vitamin C on postoperative opioid consumption and pain.[4–7] These analyses consistently conclude that vitamin C may play a beneficial role within multimodal postoperative analgesia. The majority of included trials administered a single 1–2 g dose before or during surgery. Both analyses reported statistically and clinically significant reductions in opioid requirements, with decreases of 8.0 mg (95% CI: −12.1 to −3.8) and 3.5 mg (95% CI: −5.3 to −1.8) of parenteral morphine equivalents, respectively [4, 5]. Small but consistent reductions in pain intensity were also observed, ranging from 0.65 to 0.77 points on a 10-point scale. More recent findings indicate similar benefits in orthopedic populations: in lower limb orthopedic surgery, fewer patients receiving vitamin C experienced severe postoperative pain (56%) compared with controls (72%), and pain levels decreased more rapidly in the vitamin C group [8]. In orthognathic surgery, postoperative plasma vitamin C levels dropped by 34.6%, and lower postoperative concentrations were associated with higher analgesic consumption [9].

Evidence also suggests that vitamin C may influence broader patient centered outcomes. Improved functional recovery has been reported following hip arthroplasty, and in abdominal surgery, a single preoperative 2 g oral dose was associated with higher patient satisfaction (96% vs. 64%), reduced opioid use, and lower pain intensity [10, 11]. Anti inflammatory effects have also been documented; one randomized postoperative trial found lower C reactive protein levels in patients receiving vitamin C (median 21 mg/L) compared with controls (median 42 mg/L) [12]. Beyond postoperative populations, oral vitamin C supplementation improved pain, tender joint counts, and swollen joint scores among individuals with rheumatoid arthritis [13]. Across these studies, vitamin C dosage regimens have varied considerably, from a single 25 g intravenous infusion to repeated administration of lower oral or intravenous doses over multiple days (e.g., 500 mg IV twice daily for three days; 500 mg orally every eight hours for five days; 200 mg orally three times daily for ten days; or a single 2 g oral dose) [4, 14–16].

Acute musculoskeletal (MSK) injuries presenting to the emergency department (ED), including trauma to muscles, ligaments, tendons, bones, and nerves, produce inflammation and pain levels comparable to those observed in postoperative settings [17, 18]. Management typically involves a multimodal approach that integrates pharmacologic treatments, physical therapy, activity modification, and psychological strategies [19, 20]. These injuries represent a substantial proportion of ED visits and are associated with high levels of acute pain that may persist and contribute to chronic pain development [21]. Our previous work indicates that most ED patients with traumatic MSK injuries experience moderate to severe pain, especially within the first three days after their visit [22]. Despite the biological rationale and the growing body of perioperative evidence supporting vitamin C’s analgesic potential, no studies have examined its effectiveness in managing acute pain during ED visits for traumatic injuries such as fractures, contusions, sprains, and strains.

### Study objectives

The aim of this pilot study was to assess the feasibility of conducting a randomized, placebo-controlled trial to determine the analgesic effects of vitamin C compared to placebo (lactose) during a three-day regiment in ED-discharged patients with acute musculoskeletal (MSK) injuries. The secondary aims were to obtain preliminary results on pain intensity measured by the time-weighted sum of pain-intensity difference (SPID) over a period of 72 hours (SPID72), for patients receiving vitamin C and those receiving a placebo.

## Methods

### Study Design

We conducted a double-blind, placebo-controlled, parallel-group pilot randomized trial at a single tertiary, university-affiliated trauma center in Montreal, Québec, Canada, with an annual emergency department census of approximately 60,000 visits. The study is reported in accordance with the CONSORT 2025 (Consolidated Standards of Reporting Trials) guidelines for parallel-group randomized trials [23, 24].

### Patient and Public Involvement

The study design and the pain medication diary were developed with contribution from a patient partner. The study protocol was approved by the local Research Ethics Committee. (Comité d’éthique de la recherche du CIUSSS du Nord-de-l’Île-de-Montréal).

### Participants and Recruitment

Patients with acute MSK pain for <48 hours were approached by the treating clinician (weekdays 08:00–20:00, weekends 08:00–16:00) and asked for verbal consent to meet a research assistant. Recruitment occurred from September 18, 2025, to February 4, 2026. NSAID or acetaminophen use was at the physician’s discretion. The research assistant confirmed eligibility, explained the protocol, and obtained written informed consent. Patients who declined participation were asked to provide their reason for refusal.

#### Eligibility Criteria

Patients were eligible for inclusion if they met all of the following criteria: (1) aged 18 and over; (2) treated in ED for acute MSK injury pain present for less than 48 hours (time to presentation for most acute MSK pain in our previous study) [25]; (3) Verbal numerical rating scale (VNRS) pain intensity at triage of > 3 on a 0-10 scale (at least moderate pain intensity); (4) Triaged to the ambulatory section of the ED; (5) Able to communicate in French or English.

Patients meeting any of the following criteria were excluded from the study: (1) Using vitamin C supplements currently or in the last week (half-life 8 to 20 hours [26]; (2) Active cancer; (3) Treated for any pain one hour before triage in the ED; (4) Treatment for chronic pain; (5) Unable to fill out a pain intensity diary or unavailable for follow-up; (6) Allergy to milk (lactose in the placebo) or vitamin C; (7) Treated with cyclosporine or warfarin (interaction with vitamin C); (8) Pre-existing oxalate nephropathy, liver cirrhosis or hemochromatosis.

#### Randomization Method and Blinding

At their initial visit, eligible patients were block-randomized (variable block sizes) in a 1:1 ratio to receive either 900 mg of oral vitamin C twice daily or placebo (lactose), using a centralized web-based randomization system. Because fractures are associated with higher pain intensity, randomization was stratified by fracture status (yes/no) [27, 28]. An independent pharmacist, blinded to group allocation, dispensed pre-packed, identically appearing numbered bottles of vitamin C or placebo in accordance with the centralized system.

### Study Drug

Vitamin C is an essential antioxidant nutrient required for the formation and maintenance of bones, skin, and blood vessels, not synthesized by the human body and obtained from dietary sources or over-the-counter supplements. Based on dosages used in previous studies on acute postoperative and chronic pain, reported adverse events, oral absorption and the maximum recommended daily limit for pregnant or lactating patients [29, 30], participants in the treatment arm received 900 mg of oral vitamin C twice daily (morning and evening) for three days following ED discharge, while the control arm received placebo (lactose).

### Subject Compliance Monitoring

Participants were instructed to complete a six-day diary, either electronically or on paper. Each day, they recorded whether they had taken their study capsule, their pain intensity and whether they had used any other pain medication during the day. Based on our previous work [25, 31, 32], nearly all patients can accurately understand and complete the diary and questionnaire.

### Study Procedures

After confirming eligibility and obtaining written informed consent, research assistants recorded baseline data in REDCap (Vanderbilt University, TN), hosted at Hôpital Sacré-Coeur de Montréal [33]. Collected information included a unique patient identifier, demographics (sex, gender, age, ethnicity), contact numbers, ED length of stay, final diagnosis (injury type and severity), opioids administered during the ED visit, pain intensity at triage and discharge, work-related injury status, and salary insurance coverage.

A pharmacist dispensed either the active study capsules or placebo, and patients were instructed not to start any other vitamins or natural health products during the 6-day follow-up period. The baseline questionnaire was completed with the support of a research assistant in the ED and each participant received a validated six-day electronic or paper diary. Participants were trained in person to use the diary and instructed to manage their pain with as needed acetaminophen and NSAIDs if recommended by the treating physician. Research assistants were available by phone for any questions or assistance. Real-time information on the type, quantity, and timing of all pain medications consumed, including vitamin C related to the ED visit was captured daily in the diary. Pain intensity was assessed twice daily using a 100-point plasticized visual analog scale (VASp; 0 = no pain, 100 = worst imaginable pain) provided by research assistants for both in-ED and at-home use [34]. Participants also indicated daily whether their pain had been adequately relieved (yes/no).

At the end of six days, participants were contacted by phone. They reported current pain intensity using the VASp, any additional pain medication prescriptions or clinician visits related to their initial injury, if they had any adverse event, and whether they underwent surgery related to the initial pain condition. They were also asked about use of other pain-relieving products (e.g., cannabis, alcohol, vitamins, or over-the-counter/natural products) and additional vitamin C supplements beyond the study medication. Finally, we measured their quality of life using the EQ-5D-5L questionnaire, and participants reported if and when they returned to normal activities after ED discharge. In the event of an unexpected serious adverse reaction, the research team would notify the Natural Health Products Directorate within seven days if the event was fatal or life-threatening, or within 15 days if it was not.

### Outcomes

The primary feasibility outcome was the recruitment rate. Secondary outcomes included rates of missed eligible patients, exclusions, consent, follow-up, reasons for exclusion or refusal, and treatment compliance. Compliance was defined as taking at least five of six study capsules (vitamin C or placebo). As feasibility comparator for our results, we used data from a recent pilot study that we conducted on a similar population, but only during weekdays, in which we recruited 9.2 participants per month [35]. Since the current pilot study included recruitment during weekends, feasibility was therefore defined as achieving at least 12 participants per months. We also reported the time-weighted sum of pain-intensity difference (SPID) over a period of 72 hours (SPID72), during vitamin C or placebo intake, without unblinding, describing groups as A or B.

### Sample Size

According to guidelines for designing pilot studies, ideally 30 participants should be recruited per group [36]. With two treatment arms, we aimed at recruiting a total of 60 participants.

### Statistical Analysis

Both intention-to-treat and per-protocol analyses were performed, with the latter including only participants who fully adhered to the study protocol and completed follow-up. Although this pilot study was not powered as a superiority randomized controlled trial, the per-protocol analysis was included to explore potential treatment effects, as non-compliance and crossover can reduce observed differences between groups [37]. Descriptive statistics were used to compare baseline characteristics among patients who declined participation and participants, treatment groups A and B, and participants lost to follow-up and those who completed the study. Continuous variables were summarized as means with standard deviations (SD), non-normally distributed variables as medians with interquartile ranges (IQR), and categorical variables as proportions. The same descriptive approach was applied to examine socio-demographic characteristics of each group.

SPID72 least square means differences and the corresponding 95% CI between participants in group A and group B were estimated. To determine the SPID72 we first calculated, for each of the six follow-up time points, the change in pain intensity from the baseline (baseline pain intensity – follow-up pain intensity at each time point). Then, to weigh this change, we multiplied each pain difference by the time elapsed since the previous observation. Finally, we added up all the weighted differences to obtain the SPID72 [38]. Missing pain intensity scores were imputed using k-nearest neighbors (k = 2) with a simple mean approach. SPID72 least square means differences (95% CI) were estimated using an analysis-of-covariance model (ANCOVA) adjusted for fracture status and baseline pain. Data formatting was performed using SAS version 8.3 (SAS Institute Inc., Cary, NC, USA) and statistical analyses were performed using SPSS version 26 (IBM. Somers, NY, USA).

## Results

### Study Cohort Description

Sixty participants were randomized. The mean age (SD) was 41.8 (14.2) years, 50% were female, and the mean pain intensity at triage was 7.2 (1.7) (Table 1). Compared with patients who declined participation, enrolled participants tended to be slightly older (41.8 [SD 14.2] vs 37.7 [SD 17.1]), had a higher proportion of females (50.0% vs 41.7%), higher fracture (18.3% vs 12.5%), and fewer contusions (5.0% vs 20.8%). Participants were also more likely to receive opioids during their ED stay than those who refused participation (11.7% vs 0.0%), and their ED length of stay was modestly shorter (5.3 vs 5.6 hours).

**Table 1.**
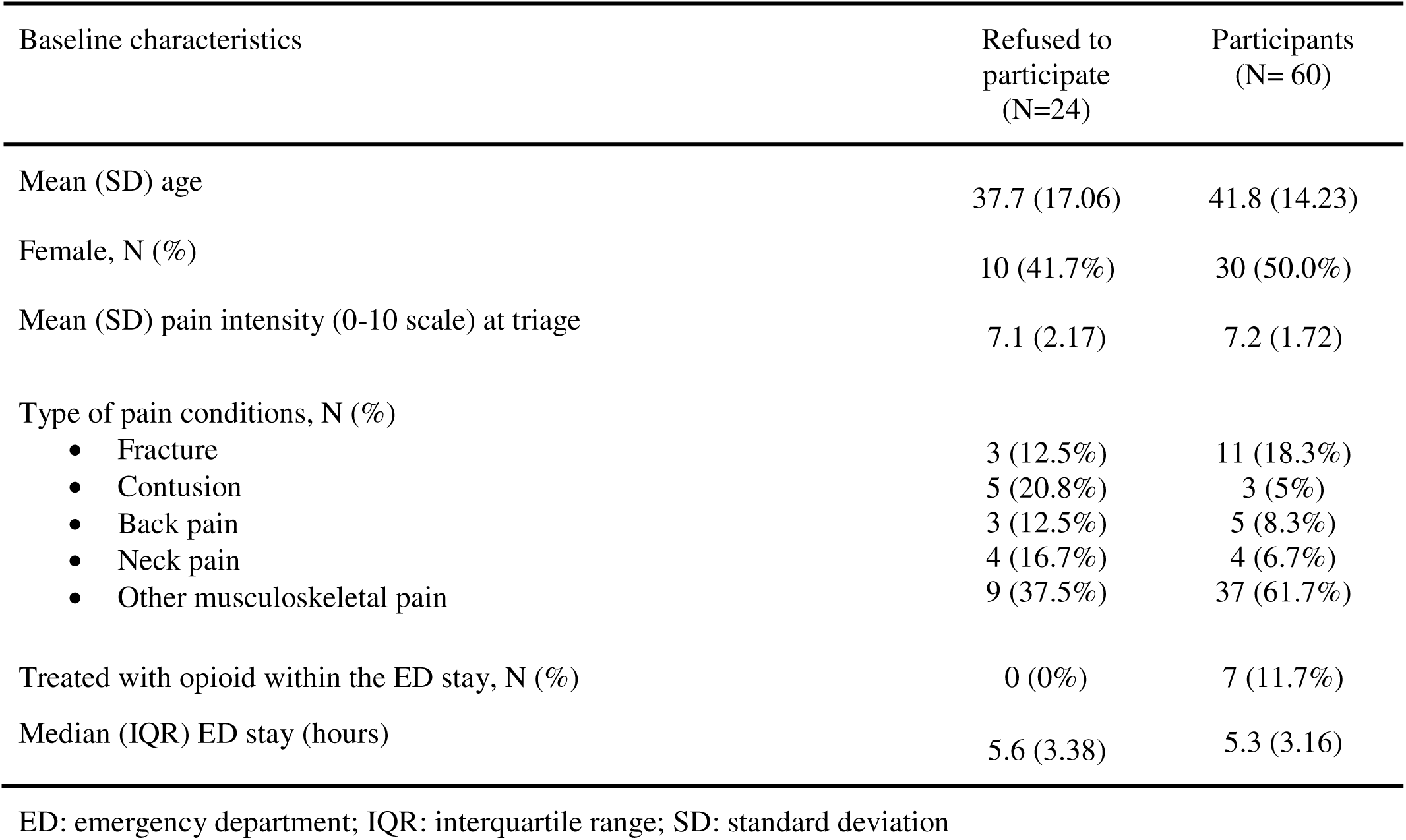
Comparison of baseline characteristics between patients who refused to participate and those who accepted to participate in the study.

Socio-demographic characteristics by treatment group are presented in Table 2. Most variables were comparable between group A and group B. Minor differences were observed in ethnicity, with a higher proportion of Black participants in group A (21.9% vs 3.6%) and a lower proportion of Arab participants (15.6% vs 32.1%) compared with group B. Opioid administration during the ED stay was more frequent in the group A than in group B (15.6% vs 7.1%).

**Table 2.** Comparison of baseline characteristics between group A and group B participants.

| Baseline characteristics and analgesic prescription | Group A (N= 32) | Group B (N= 28) |
| --- | --- | --- |
| Mean (SD) age | 41.1 (13.99) | 42.6 (14.72) |
| Female, N (%) | 14 (43.8%) | 16 (57.1%) |
| Ethnicity, N (%) |  |  |
| • White | 12 (37.5%) | 14 (50%) |
| • Arab | 5 (15.6%) | 9 (32.1%) |
| • Black | 7 (21.9%) | 1 (3.6%) |
| • Other | 8 (25.0%) | 4 (14.3%) |
| Education level, N (%) |  |  |
| • Primary/High school | 8 (25.0%) | 7 (25%) |
| • College/University | 24 (75.0%) | 20 (71.4%) |
| • Other | 0 (0.0%) | 1 (3.6%) |
| Smoker, N (%) | 10 (31.3%) | 8 (28.6%) |
| Mean (SD) pain intensity (0-10 scale) at triage | 6.9 (1.97) | 7.5 (1.33) |
| Mean (SD) pain intensity (0-10 scale) at ED discharge | 5.0 (1.9) | 5.2 (2.1) |
| Type of pain conditions, N (%) |  |  |
| • Fracture | 6 (18.8%) | 5 (17.9%) |
| • Contusion | 1 (3.1%) | 2 (7.1%) |
| • Back pain | 1 (3.1%) | 4 (14.3%) |
| • Neck pain | 3 (9.4%) | 1 (3.6%) |
| • Other musculoskeletal pain | 21 (65.6%) | 16 (57.1%) |
| Median (IQR) ED stay (hours) | 5.7 (4.66) | 5.1 (2.25) |
| Treated with opioid within ED stay, N (%) | 5 (15.6%) | 2 (7.1%) |

ED: emergency department; IQR: interquartile range; SD: standard deviation

### Feasibility Outcomes

During the recruitment period, 147 patients were screened, including 68 who were missed by treating physicians. Of those screened, 63 patients (42.9%) were excluded (Figure 1), most commonly because they declined to answer screening questions, were unable to complete a diary or participate in follow-up, or reported chronic pain. Among the 84 eligible patients, 60 were randomized (71.4%) and 24 (28.6%) declined participation, primarily due to lack of interest, time constraints, reluctance to add or mix medications, or concerns about potential side effects.

**Figure 1.**
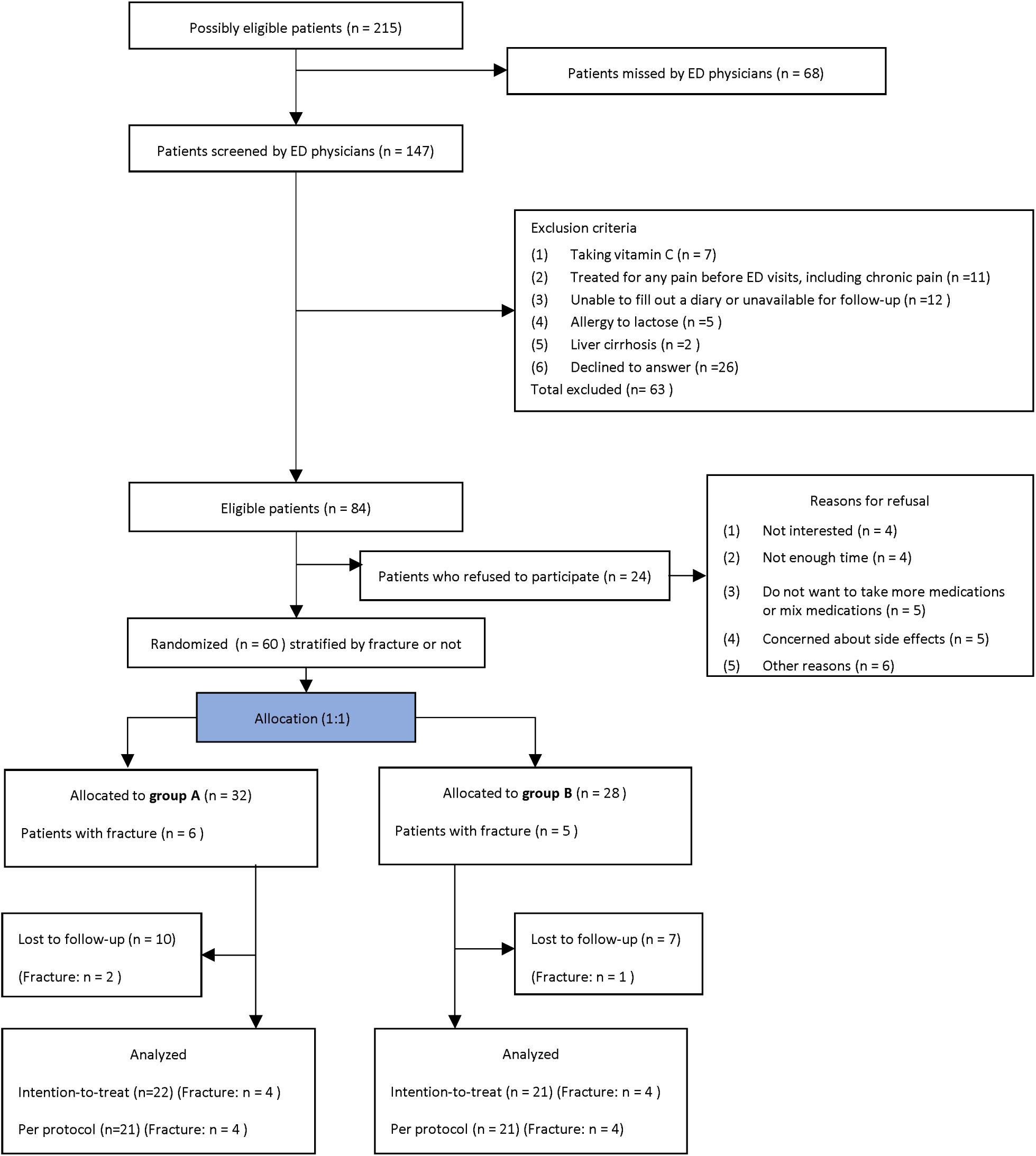
Flow chart of patients’ enrollment in the study. Legend: ED: emergency department

Sixty participants were randomized: 32 to group A and 28 to the group B. Fractures were present in six participants in the group A and five in the group B. The recruitment rate was 13.0 participants per month. Overall, 43 participants (71.7%) completed follow-up. Lost to follow-up rate differed between participants with electronic diary (n=7; 16.7%) and participants with paper diary (n=10; 55.6%). This discrepancy may be partly attributable to the Canada Post strike that took place during the study, as participants were required to mail their paper diaries.

Baseline characteristics of participants lost to follow-up were generally similar to those who completed follow-up, although the group who completed follow-up seemed younger (Table 3). Contusions and neck pain were more common, and back pain less common, in those lost to follow-up. Treatment adherence was high at 97.6%, with only one participant in the group A classified as non-compliant.

**Table 3.**
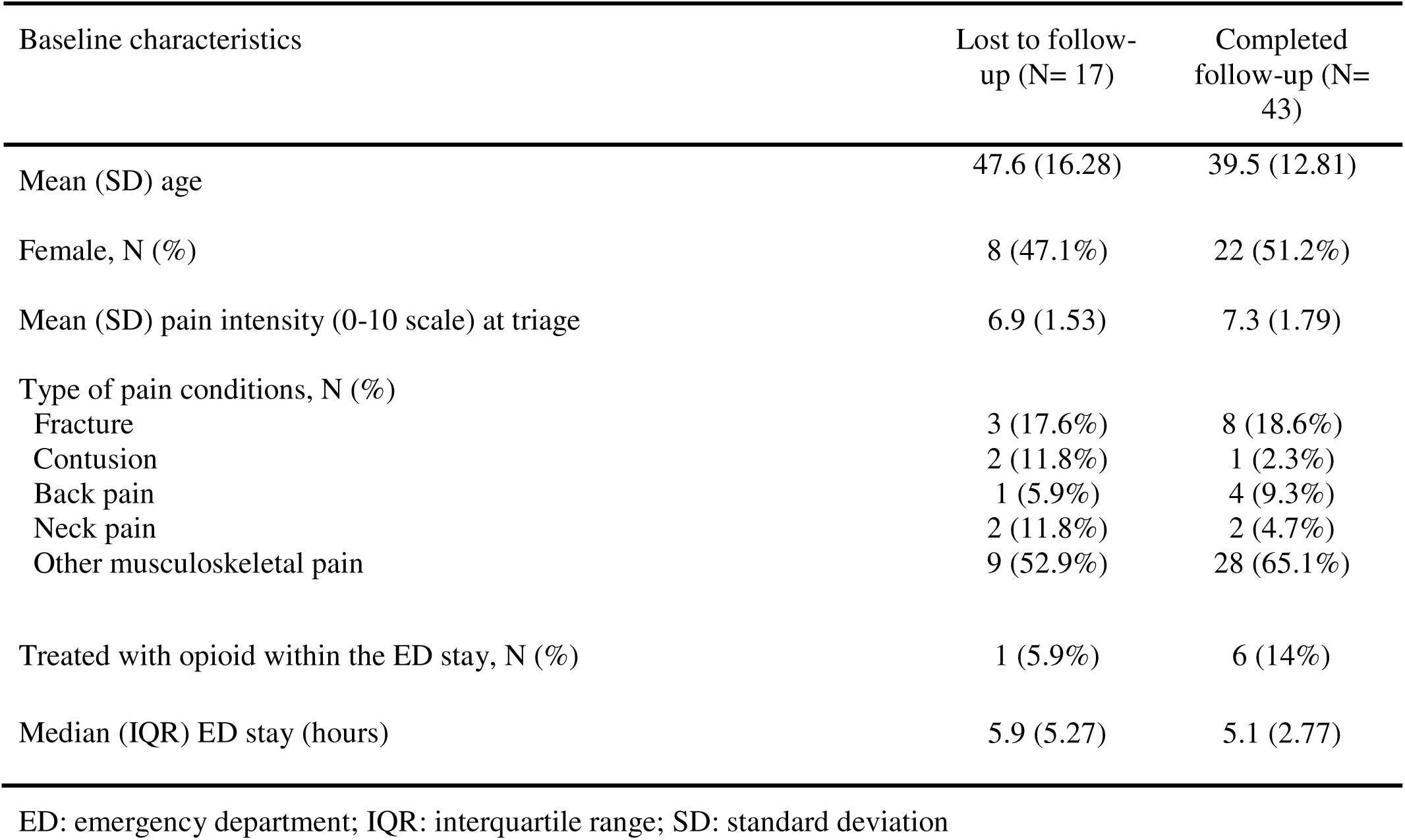
Comparison of baseline characteristics between participants who were lost to follow-up and those who completed follow-up.

### Sum of pain intensity difference during the first three days

SPID72 least square means differences (95% CI) for both groups and for intention-to-treat and per-protocol analyses are presented in Table 4. The mean values between group A and group B seemed comparable.

**Table 4.** SPID72 least square means differences between participants in group A and group B for intention-to-treat and per-protocol analysis.

|  | Group A - Group B |
| --- | --- |
| Intention to treat analysis | (N= 43) |
| LSM difference SPID72 (95% CI) | 348.7 (-698.9, 1396.4) |
| Per protocol analysis | (N= 42) |
| LSM difference SPID72 (95% CI) | 357.6 (-709.67, 1424.82) |

CI: confidence interval; LSM: least square means; SPID72: sum of pain intensity differences over 72 hours

## Discussion

Findings from this pilot study support the feasibility of conducting a full-scale randomized, placebo-controlled trial to evaluate the analgesic effect of vitamin C compared with placebo over a six-day follow-up period in ED-discharged patients with acute musculoskeletal injuries. While a high number of eligible patients were missed, the study still met its recruitment feasibility targets. This recruitment gap likely reflects clinician confusion arising from the concurrent conduct of another study with overlapping but distinct eligibility criteria. Minimizing the concurrent running of similar studies, improving clinician awareness of eligibility criteria differences across active studies, and approaching patients before evaluation by an ED clinician, such as immediately following triage, may collectively reduce missed recruitment opportunities and improve enrollment. Notably, in a prior multicenter study involving a larger cohort and no concurrent similar studies, we observed a substantially lower proportion of missed eligible patients (15.6%). [39].

The observed consent rate (71.4%) was consistent with previously reported randomized controlled trial (RCT) benchmarks (median (IQR): 72% (50%–88%)).[40] Participants with shorter ED stays and those receiving opioids appeared more willing to enroll. This aligns with patients declining participation because of time constraints, or reluctance to take additional medications. Since patients are recruited when they are being discharged from the ED, providing effective analgesia and timely healthcare in the ED or recruiting earlier during ED stay might help reduce the refusal rate in future RCT. Also, to address the lack of interest in the study and to reduce concerns about potential side effects of the vitamin C, providing more information on the waiting room monitors and visual infographic leaflets aids could help.

Baseline characteristics were generally comparable between group A and group B, with only minor differences in ethnicity. However, a small pilot study cannot be used to inform randomization performance [36]. Our retention rate of 71.7% is lower than the median (IQR) reported in previously reported RCT benchmarks 88% (80%-97%) [40]. Loss to follow up was higher among participants using paper diaries than among those using electronic diaries (55.6% vs. 16.7%). A Canada Post strike during the study may partly explain this difference: of nine paper diaries not received, two participants confirmed to our research assistant that they had mailed their diaries, which nevertheless were not delivered. Moreover, participants lost to follow-up appear to have a longer length of stay, slightly lower pain intensity at triage, and to be treated less often with opioids during their ED stay compared to those who completed the study. It is likely that these participants experienced little pain during follow-up and therefore had less interest in keeping up with the study requirements. Using different mailing systems and promoting the electronic diary version could improve the retention rate. Compliance with study capsules was high among participants with follow-up data, with only one of 43 participants failing to comply. This suggests that participants who completed follow-up understood the importance of adhering to the study treatment, contributing to more accurate data. Finally, given the pilot design and small sample size, no conclusions regarding the vitamin c effect compared with the placebo group can be drawn.

### Limitations

This pilot RCT was not powered to detect clinical differences between groups, and any comparative findings should be interpreted cautiously. Feasibility was assessed in a single academic center, which may limit generalizability to other settings. A high number of missed eligible participants likely resulted from clinician confusion related to a concurrent study with overlapping eligibility criteria.

Despite these challenges, the target sample size was achieved within a reasonable timeline, supporting the feasibility of a future large-scale trial while highlighting areas for procedural improvement.

### Strategies to Optimize Recruitment and Retention

Several targeted measures could be implemented to enhance participant recruitment and minimize loss to follow-up. Improved communication with clinicians regarding eligibility criteria, particularly differences from currently active studies, combined with earlier patient identification during the ED visit (e.g., immediately following triage) would reduce missed recruitment opportunities. Stationing a dedicated research assistant in the emergency department to prospectively identify and approach eligible patients prior to clinician evaluation could further strengthen enrollment.

To reduce loss to follow-up, the consent process should be used strategically to emphasize the importance of completing all follow-up procedures, thereby sensitizing participants to the importance of remaining engaged throughout the study. Retention could be further supported through additional text message reminders during the diary completion period, supplemented by follow-up phone calls for participants using paper diaries to prompt the timely return of completed forms. All participants should receive a reminder card specifying the date and time of their scheduled follow-up call, with an automated text reminder sent the preceding day.

## Conclusion

This pilot study supports the feasibility of a larger randomized controlled trial on the analgesic properties of vitamin C for acute MSK injured ED patients. Strategies to reduce the missed patients and lost to follow-up rates are proposed.

## Data Availability

All data produced in the present study are available upon reasonable request to the authors

## Acknowledgments

We would like to thank Martin Marquis for English language editing.

## Contributors

RD conceived the study and obtained research funding. RD, DW, VH, CA, JP, SB, ME, DR, and AC contributed to the final protocol and data interpretation. MK was responsible for data management and statistical analysis. RD and MK drafted the manuscript. DW, VH, JL, CA, ME, JP, DR, SB, and AC contributed substantially to its revision. All authors approved the final manuscript as submitted and have agreed to be accountable for all aspects of the work.

Award/Grant number: N/A.

## Role of the Funder/Sponsor

The funders had no role in study design, data collection and analysis, decision to publish, or preparation of the manuscript.

## Competing interests

None declared.

## Provenance and peer review

Not commissioned; externally peer reviewed.

## Notes

### Competing Interest Statement

The authors have declared no competing interest.

### Clinical Trial

NCT06306183

### Funding Statement

Chaire Dr Sadok Besrour

### Author Declarations

The study protocol was approved by the local Research Ethics Committee. (Comite d'ethique de la recherche du CIUSSS du Nord-de-l'Ile-de-Montreal).

